# Cellular aging is accelerated in the malignant clone of myeloproliferative neoplasms

**DOI:** 10.1101/2023.09.12.23295349

**Authors:** Margherita Vieri, Vithurithra Tharmapalan, Milena Kalmer, Julian Baumeister, Miloš Nikolić, Matthis Schnitker, Martin Kirschner, Niclas Flosdorf, Marcelo A. S. de Toledo, Martin Zenke, Steffen Koschmieder, Tim H. Brümmendorf, Fabian Beier, Wolfgang Wagner

## Abstract

Myeloproliferative neoplasms (MPNs) are caused by somatic driver mutations, such as *JAK2*^V617F^, which might also affect cellular aging and senescence. Here, we analyzed the heterogeneity of aging in MPN patients and if this can be used to specifically target malignant cells. The mean epigenetic age was significantly accelerated in 129 MPN patients across all disease-entities, whereas premature telomere attrition was particularly observed in primary myelofibrosis. Overall, accelerated cellular aging correlated with *JAK2*^V617F^ allele frequency and was more pronounced in colony forming cells with *JAK2*^V617F^ as compared to *JAK2* wild-type colonies. *JAK2*^V617F^ mutation did not evoke clear acceleration of aging in syngeneic iPSC models upon short-term hematopoietic differentiation. On the other hand, a murine *Jak2*^V617F^ model revealed epigenetic age-acceleration that therefore appears as sequel of disease progression. To investigate if the malignant clone might be targeted, we tested eight senolytic compounds, of which JQ1 and piperlongumine showed a reduction in allele burden and an increase in telomere length. Notably, treatment with the telomerase inhibitor BIBR-1532 reduced mutated colonies, particularly in patients with preexisting short telomeres. Our results indicate that cellular aging is accelerated in malignant MPN clones and this can provide a target for treatment with senolytic drugs or telomerase inhibitors.

## Introduction

Myeloproliferative neoplasms are clonal hematopoietic stem cell disorders characterized by the excessive production of differentiated blood cells. The majority of MPN patients carry a point mutation in Janus kinase 2 (*JAK2*) at the position 617 (valine to phenylalanine, V617F), which is observed in 95% of patients with polycythemia vera (PV), and 50-60% with essential thrombocythemia (ET) and primary myelofibrosis (PMF) (1). Other driver mutations, which are mutually exclusive, affect the genes calreticulin (*CALR)* and myeloproliferative leukemia protein (*MPL)* (2–4). Somatic mutations in MPN may not only drive proliferation - they could also accelerate replicative and cellular aging as well as genetic instability, potentially contributing to disease progression (5).

Telomere attrition represents a common denominator of cellular aging and its impact has been studied extensively in BCR-ABL-positive chronic myeloid leukemia (CML). Telomere length (TL) was found significantly shortened in the malignant CML stem cell clone (6). Furthermore, the degree of telomere shortening correlated with leukemic involvement of the hematopoietic stem cell compartment (7), duration and stage of the disease, as well as prognosis and response to treatment (8). These findings resulted in a model of phase transition for CML, where TL served as a marker of progression (6, 8, 9). In analogy, there is evidence that TL is also severely reduced in BCR-ABL-negative MPN (10) and telomerase activity was found to be increased in almost all MPN subgroups (11).. Thus, there is evidence that cellular aging is accelerated in MPN, but little is known how this is related to different entities, driver mutations, and if it specifically affects the malignant clone.

Aging is also reflected by epigenetic modifications, such as changes in DNA methylation (DNAm) (12). It has been shown that epigenetic age predictions often reveal clear offsets in hematopoietic malignancies due to the prevailing DNAm pattern of the malignant clone (13). Despite this aberration, age-associated DNAm changes seem to be coordinately regulated and correlate with clinical parameters and overall survival in several types of cancer (13). So far, little is known about the heterogeneity in epigenetic aging within a given patient and whether this is specifically accelerated in the malignant clone.

Senolytic drugs raised high hopes to specifically target prematurely aged cells that show signs of senescence and to thereby rejuvenate tissues (14). Similarly, senolytic compounds might also selectively target malignant cells if they reveal molecular signs of accelerated cellular aging. In line with this concept, imetelstat, a telomerase inhibitor (TI), has shown improved outcomes in clinical trials in MPN patients (15–17). Nevertheless, whether telomere attrition or epigenetic changes specifically occur in the malignant MPN clone and whether TI or other senolytic treatments target the malignant clone remains to be demonstrated. In this study, we have therefore systematically evaluated how cellular aging is affected across different MPN entities and if it can be utilized to specifically target the malignant clone.

## Materials and Methods

### Blood samples

Peripheral blood samples of 154 patients diagnosed with MPN were used. Blood samples of 134 healthy donors were used for age-adaptation of TL measured via flow-FISH (18). Furthermore, a separate cohort of 128 healthy controls was used for the measurement of epigenetic age deviation (19). All samples were obtained after informed and written consent in accordance with the Declaration of Helsinki and the research was specifically approved by the local ethics committee of RWTH Aachen University (EK 041/15, EK 206/09 and EK 127/12).

### Telomere length analysis

Cryopreserved whole blood cells after red blood cell depletion were used for the flow- fluorescent in-situ hybridization (flow-FISH) analysis of TL in lymphocytes and granulocytes (18). Alternatively, TL of colonies derived from CFUs, CD34^+^ and PBMC cell fractions were analyzed by telomere-PCR (TEL-PCR) (20).

### Epigenetic age-prediction

Genomic DNA was isolated from cryopreserved whole blood cells after red blood cells depletion, bisulfite converted, and then used for targeted bisulfite amplicon sequencing (BA- seq) and pyrosequencing. Three age-associated CG dinucleotides (CpG sites) that are associated with the coiled-coil domain-containing protein 102B (*CCDC102B*), four and a half LIM domains protein 2 (*FHL2*) and phosphodiesterase 4C (*PDE4C*) (19) were analyzed. PCR conditions are summarized in Supplemental Table S1.

### Colony forming unit assay

Peripheral blood mononuclear cells (PBMCs) of MPN and seven healthy donors were isolated by gradient centrifugation with Pancoll (Pan Biotech, Aidenbach, Germany) and 1x10^6^ cells per condition were transferred on a semisolid medium to perform CFU (21). After 14 days, colonies were counted and classified. To determine the mutation status of *JAK2* and *CALR* in single colonies, PCRs were performed for *JAK2*^V617F^, *CALR*^ins5^ and *CALR*^del52^ (Supplemental Table S2) (21).

### Drug testing

Peripheral blood mononuclear cells of MPN patients were cultured for three days with nine different compounds: nutlin3a, ABT263, piperlongumine, S63845, RG7112, BIBR-1532, JQ1, dasatinib combined with quercetin, and AMG232. Epigenetic age, TL and *JAK2*^V617F^ frequency were measured to determine the effect of senolytics.

### Additional methods

Additional methods for sorting of CD34^+^ cells, mutation burden measurement, senescence associated beta-galactosidase [SA-ß-gal] assay, analysis of microarray data, induced pluripotent stem cell (iPSC) lines with and without *JAK2*^V617F^ mutation and their hematopoietic differentiation, the *Jak2*^V617F^ mouse model, and statistics are provided in the supplemental methods. Furthermore, more detailed descriptions are provided there for TL measurements, epigenetic age prediction and drug testing.

## Results

### Epigenetic age-predictions and telomere length in MPN samples

Epigenetic age predictions were initially validated with blood samples of 128 healthy donors (74 males and 54 females) aged 18-74 years, showing a mean age deviation (MAD) of 0.8 years, and a correlation with chronological age of R^2^ = 0.86 (Figure 1a). In contrast, epigenetic age-predictions of 129 MPN patients showed much higher offsets, particularly in the advanced MPN entity myelofibrosis (ET: n = 43, MAD = 6.9 years; PV: n = 39, MAD = 9.8 years; PMF: n = 35, MAD = 12.9 years; post-ET/PV MF: n = 12, MAD = 10.3 years; Figure 1b). Overall, epigenetic aging was rather increased, resulting in a positive delta-age (predicted age – chronological age; Figure 1c).

**Figure 1.**
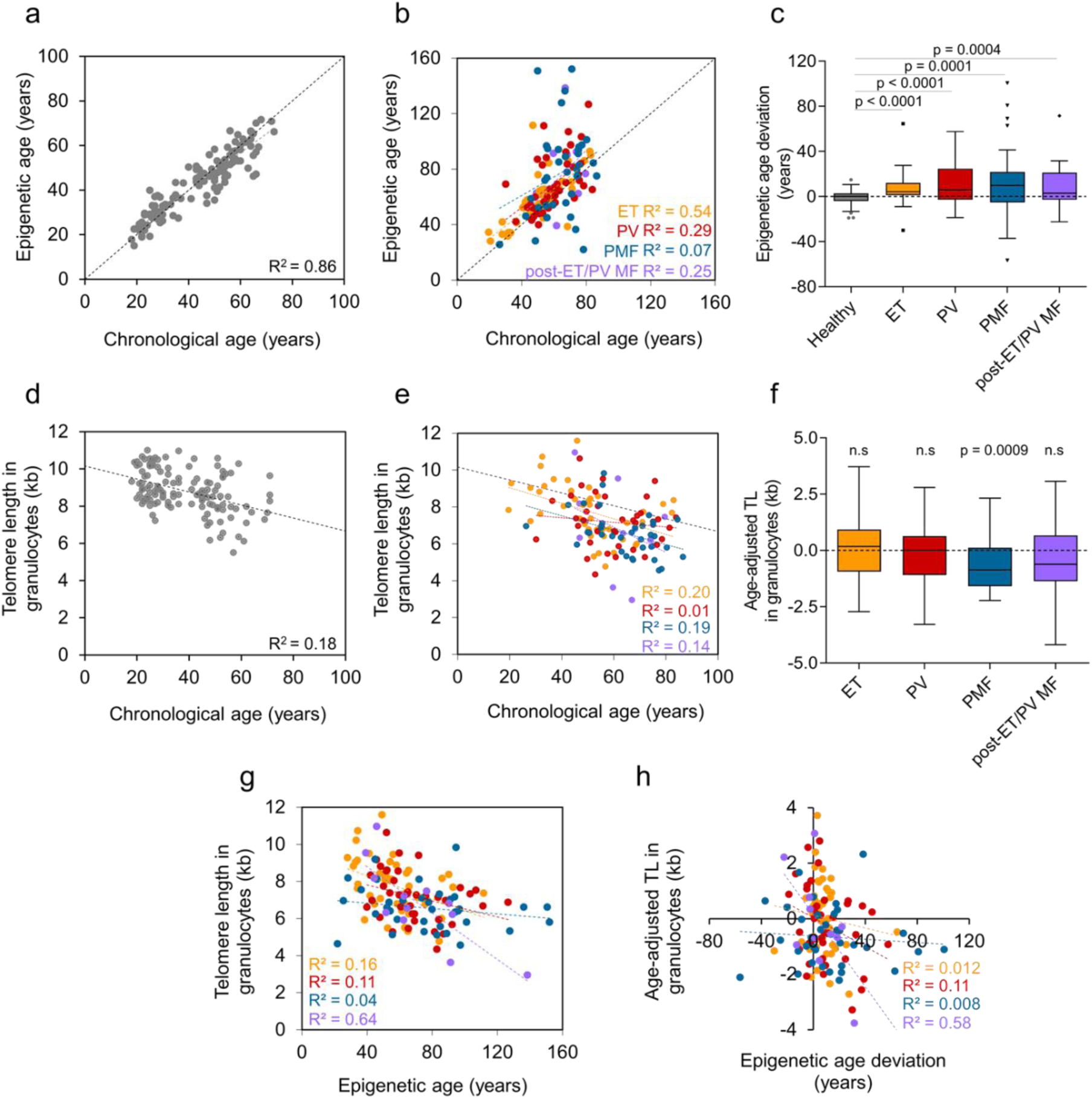
Cellular aging is progressively accelerated in MPN entities. **a)** Correlation of chronological age and epigenetic age predictions by bisulfite barcoded amplicon sequencing (BA-seq) of three CpGs in healthy donors (n = 128). **b)** Correlation of chronological and epigenetic age in MPN patients (n = 129; PV = polycythemia vera, ET = essential thrombocythemia, PMF = primary myelofibrosis). **c)** Epigenetic age deviation in different MPN entities compared to healthy donors. Unpaired t-test was used to assess statistical significance. **d)** Telomere length (TL, in kb) was measured in granulocytes via flow-FISH in healthy donors, as described before (n = 134). **e)** In analogy, we analyzed TL in blood of MPN patients (n = 129). **f)** Age-adapted TL in granulocytes in different MPN entities. One-sample t-test was used to calculate statistical significance. **g)** Correlation of TL and epigenetic age in different MPN entities. **h)** Correlation of epigenetic age deviation with age-adjusted TL in granulocytes in MPN entities.

In addition, flow-FISH analysis of telomere length in granulocytes revealed a clear association with chronological age in 134 healthy donors, albeit the correlation was lower than for epigenetic age-predictions, as demonstrated before (18) (R^2^ = 0.18; Figure 1d). In the 129 MPN samples, the distribution was overall similar (Figure 1e), while a significant telomere attrition was only observed for the PMF samples (p = 0.0009; Figure 1f). Expectedly, no relevant deviation of TL from healthy controls was observed in the lymphocyte compartment of all three MPN subgroups (Supplemental Figure S1). Since the disease-initiating cells of MPN are considered to be among the CD34^+^ fraction, we have exemplarily compared TL of 5 MPN patients in peripheral blood (PB)-derived CD34^+^ cells *versus* PBMC, but there were no significant differences (Supplemental Figure S2). Overall, there was only a moderate correlation between absolute TL and epigenetic age predictions (Figure 1g) as well as between epigenetic age-deviation and age-adjusted TL (Figure 1h), indicating that both parameters for cellular aging reflect independent biological features of cellular aging.

Next, we analyzed if these measures for cellular aging were associated with specific somatic mutations using a next-generation sequencing panel of 32 genes. Epigenetic age deviation showed a significant increase in patients with *JAK2^V617F^* (p < 0.001) and *CALR* mutations (p = 0.01; Figure 2a), and age-adjusted TL in granulocytes showed significantly accelerated shortening in samples with mutation in *CALR* (p = 0.025) and *ASXL1* (p = 0.004; Figure 2b). Notably, in patients carrying the *JAK2*^V617F^ mutation, there was a significant association of epigenetic age acceleration (p = 0.0031; Figure 2c) as well as telomere attrition (p < 0.0001; Figure 2d) with mutational burden if all MPN subgroups were combined. Despite the smaller sample sizes, the results were also significant for epigenetic age acceleration in PV (p = 0.0060) and for TL in PV (p = 0.0144), ET (p = 0.0257) and post ET/PV -MF (p = 0.0418). In summary - and in line with previous studies in CML (7) - cellular aging seems to be generally accelerated in MPN.

**Figure 2.**
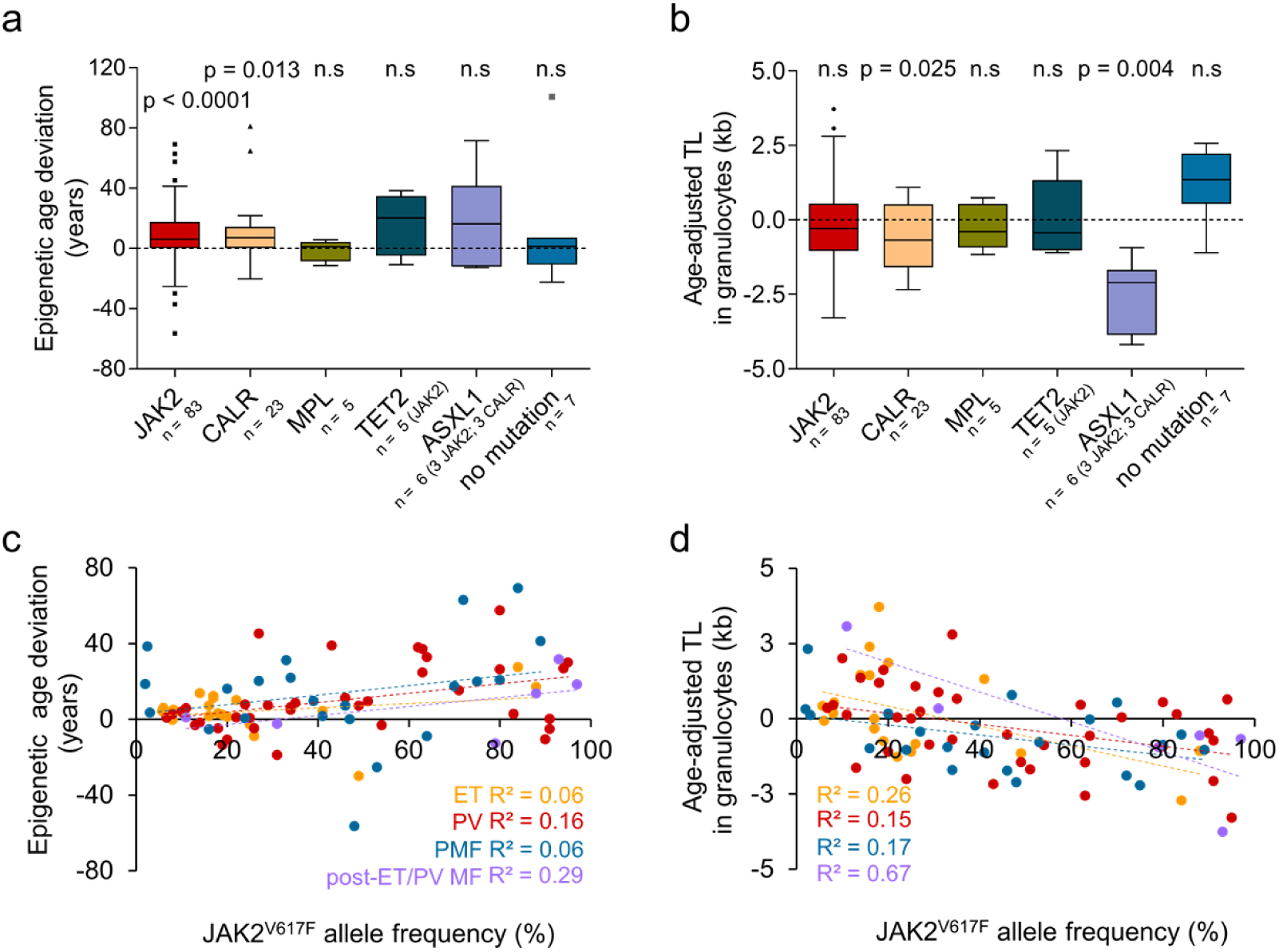
Cellular aging by mutations and correlations with JAK2V617F burden. **a)** Epigenetic age deviation in MPN carrying different driver mutations. Unpaired Welch’s t-test was used to assess statistical significance. **b)** Age-adapted TL in granulocytes in MPN carrying different driver mutations. One-sample t-test was used to calculate statistical significance. **c)** Correlation of epigenetic age deviation and JAK2^V617F^ allele burden in different MPN entities. **d)** Correlation of age-adjusted TL and JAK2^V617F^ allele burden.

### Heterogeneity of epigenetic age based on single-read predictions

It has been speculated that epigenetic age-deviations in malignancies are due to altered DNA methylation patterns of the malignant clone, rather than the residual compartment of non-mutated cells. To address this question, we investigated the individual amplicon sequencing reads. Each of the three age-associated regions capture several neighboring CpG sites within the amplicons (*PDE4C*, n = 26; *FHL2*, n = 18; and *CCDC102B*, n = 6). It might be anticipated that these neighboring sites are coherently modified in the malignant cells. In contrast, the neighboring CpGs seem to be regulated independently (Figure 3a-b; Supplemental Figure S3), as recently also observed for healthy samples (19). To gain insight into the heterogeneity of epigenetic aging, we performed single-read predictions, which are based on a probabilistic approach for a given sequel of methylated and non-methylated CpGs on a DNA strand (19). For healthy samples, such single read-predictions followed a more homogeneous pattern than in MPN patients, indicating that epigenetic aging is more heterogeneous in MPN (Figure 3c- d; Supplemental Figure S3).

**Figure 3.**
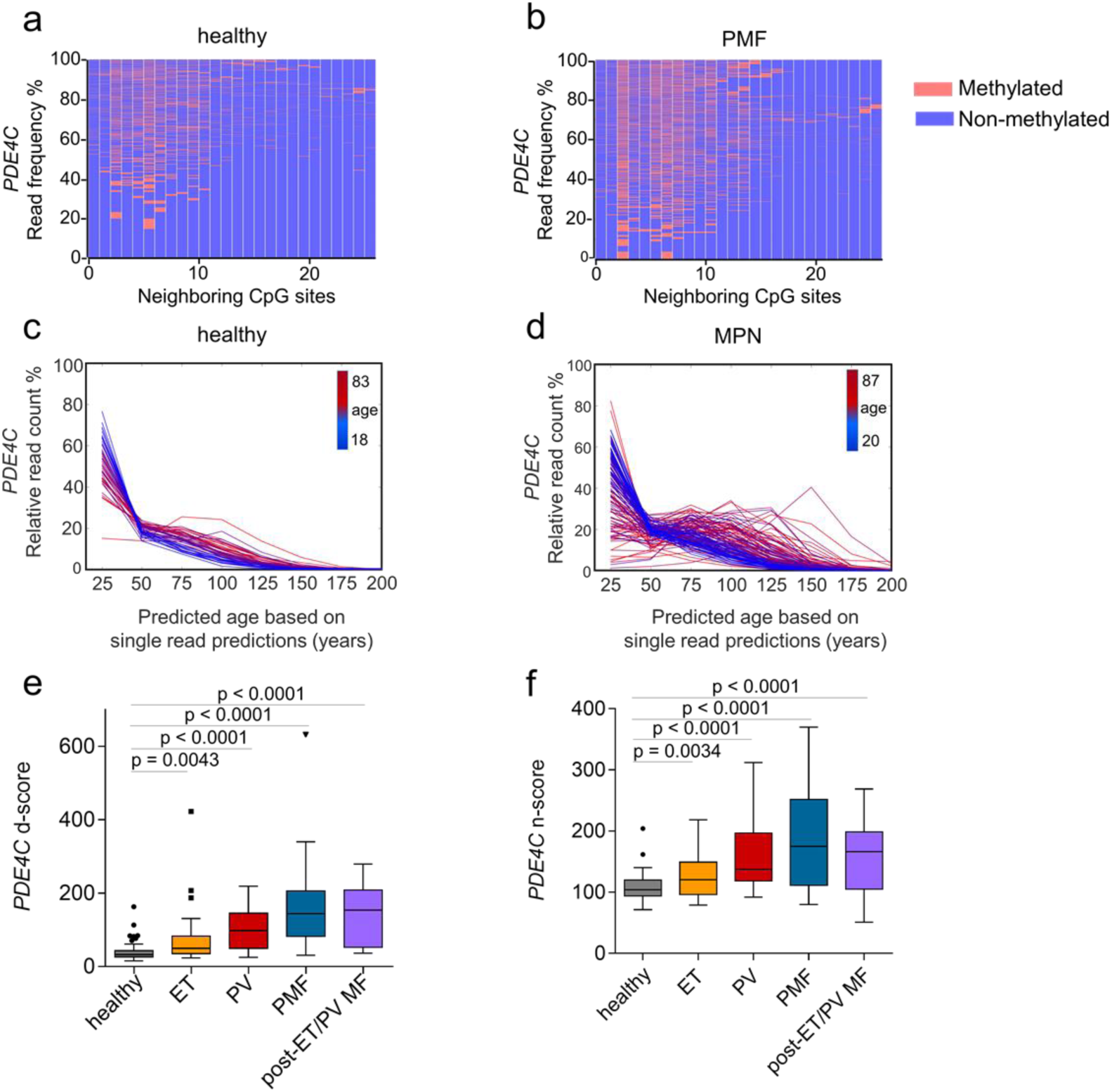
**Heterogeneity of age-associated DNA methylation in MPN.** Heat map to exemplarily depict frequencies of DNAm patterns within the neighboring CpGs of the *PDE4C* amplicon in BA-seq data of **a)** a healthy donor, and **b)** a PMF patient of the same age. The DNAm patterns in *PDE4C* were subsequently used for single read prediction with our probabilistic approach (19). The relative percentages for age-predictions of individual reads are provided for **c)** healthy donors of different ages (n = 128; color code depicts donor age), and **d)** of MPN patients (n = 129). Within MPN samples a higher heterogeneity of epigenetic age was observed, particularly in the elderly. **e)** The DNAm at the neighboring CpGs within the *PDE4C* amplicon was compared to age- adjusted levels in healthy blood samples. The d-score (22) demonstrates clear demarcation of MPN and control samples. **f)** Aberrant DNAm at age-associated CpGs often shows non-coherent DNAm levels at neighboring CpGs. This is exemplified by the n-score (22) for the *PDE4C* amplicon, which is consistently higher in MPN as compared to healthy controls. Unpaired t-test was used to assess statistical significance.

To better understand the variations in DNA methylation levels at neighboring CpGs, we utilized two alternative scoring systems, as described in our previous work (22): the d-score and the n-score. The d-score reflects how much a DNAm patterns in an MPN sample varies from a healthy control, taking all neighboring sites into account. For the 26 CpGs of *PDE4C* the d-score was significantly increased for all MPN entities as compared to healthy controls (Figure 3e). The n-score is calculated by the absolute difference between neighboring CpG sites and thereby reflects how much the DNAm levels vary within the *PDE4C* amplicons. The n-scores were also significantly increased in MPN samples, especially in PMF (Figure 3f). Overall, these data demonstrate that age-associated DNAm at neighboring CpGs is heterogeneous even within the malignant clone.

### Cellular aging in colony forming units

To further investigate cellular aging in the malignant clone of patients with MPN, we analyzed single cell derived CFUs. Epigenetic age predictions were exemplarily performed for 5 wild type (WT) and 5 *JAK2*^V617F^ colonies for an individual patient per MPN sub-entity. There was some variation between individual colonies, but overall, the epigenetic age predictions were consistently higher for the mutated clones (Figure 4a). This is in line with the correlation of epigenetic age-acceleration with *JAK2*^V617F^ mutation in the analysis conducted on the PBMCs. Notably, in contrast to the BA-seq DNAm patterns in the bulk analysis, the individual colonies revealed prevalent and colony-specific DNAm patterns (Figure 4b; Supplemental Figure S4). This demonstrates that the clonal CFUs capture a specific DNAm pattern.

**Figure 4.**
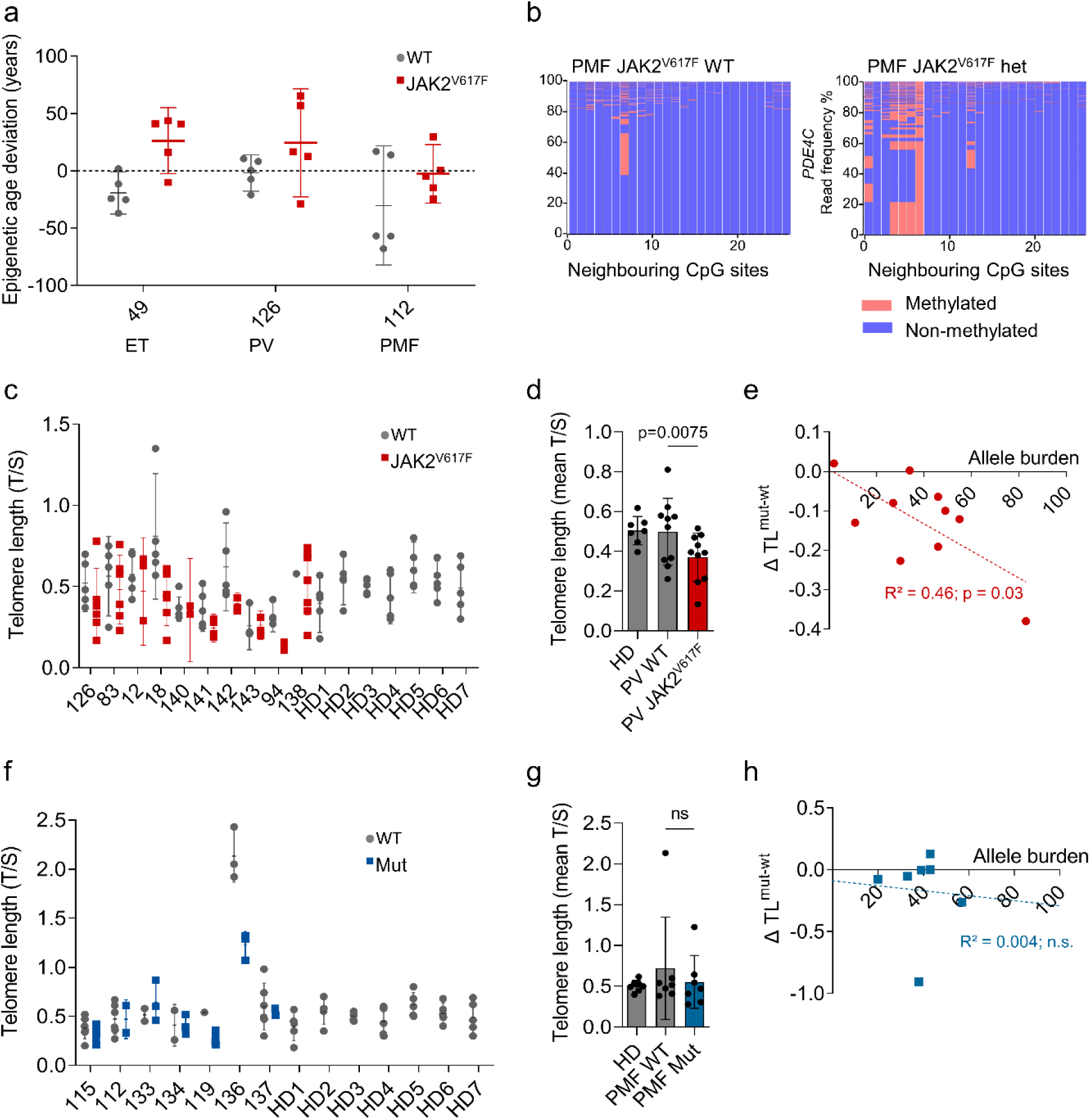
Cellular aging is accelerated in colony forming units with JAK2V617F. **a)** Epigenetic age predictions were performed in CFUs without (n = 5) and with *JAK2*^V617F^ (n = 5) in three different patients affected by either ET, PV, or PMF. **b)** Heat maps to exemplarily depict frequencies of DNAm patterns within the neighboring CpGs of the *PDE4C* amplicon in BA-seq data of a wild-type (WT) colony and a *JAK2*^V617F^ heterozygous colony from a PMF patient. **c)** Telomere length analysis (TEL-PCR) in single colonies derived from ten PV patients and 7 healthy donors (HD). For each patient, up to ten colonies were analyzed also for *JAK2*^V617F^ genotype. **d)** Mean difference in TL between WT and *JAK2*^V617F^ colonies of each individual PV patient (paired t-student test). **e)** Correlation between this mean difference in TL with the initial *JAK2*^V617F^ allele burden of the patients. **f)** In analogy, TL was analyzed in up to then single colonies derived from seven PMF patients and the same 7 healthy donors. **g)** The colonies were again classified in healthy, WT and *JAK2*^V617F^ mutated and the mean difference in TL is depicted (paired t-student test). **h)** In the PMF patients, there was no clear correlation between the initial *JAK2*^V617F^ allele burden and the discrepancy in TL between mutated and non-mutated colonies.

In analogy, we analyzed TL in individual *JAK2*^V617F^ and *JAK2*^WT^ colonies with TEL-PCR in 10 PV patients and 7 healthy donors (Figure 4c, d). Overall, the mean TL of colonies with *JAK2*^V617F^ mutation was significantly shorter than in WT colonies (p = 0.0075). The degree of telomere shortening in *JAK2*^V617F^ colonies (ΔTL^mut-WT^) was found to correlate significantly with the patient’s allele burden (Figure 4e). However, these associations were not clearly observed in 7 PMF patients (Figure 4f-h).

### Impact of genetic perturbation of *JAK2*^V617F^ mutation on cellular aging

To understand if the *JAK2*^V617F^ mutation directly affects cellular aging parameters we used previously established syngeneic induced pluripotent stem cell (iPSC) models (23). Corresponding iPSC lines were generated from three MPN patients with WT *JAK2*, heterozygous and homozygous *JAK2*^V617F^ mutations. These clones were then differentiated toward the hematopoietic lineage (Figure 5a) and they were previously shown to recapitulate the pathognomonic skewed megakaryocytic and erythroid differentiation of MPN patients (23). Here, we focused particularly on the cellular aging in the iPSC-lines and iPSC-derived hematopoietic cells. To estimate epigenetic age, we used two commonly used multi-tissue epigenetic predictors: the Horvath epigenetic clock (24) and Horvath Skin and Blood clock (25). In fact, there was a tendency towards increased epigenetic age in iPSC lines with *JAK2*^V617F^ mutation (Figure 5b), but the predictions were overall very close to 0 years, as generally observed upon reprogramming (26). Furthermore, TL analysis did not reveal consistent telomere shortening in the *JAK2* mutant clones (Figure 5c). Thus, the impact of *JAK2*^V617F^ on acceleration of cellular aging appears to be overall negligible upon short-term differentiation in the iPSC model.

**Figure 5.**
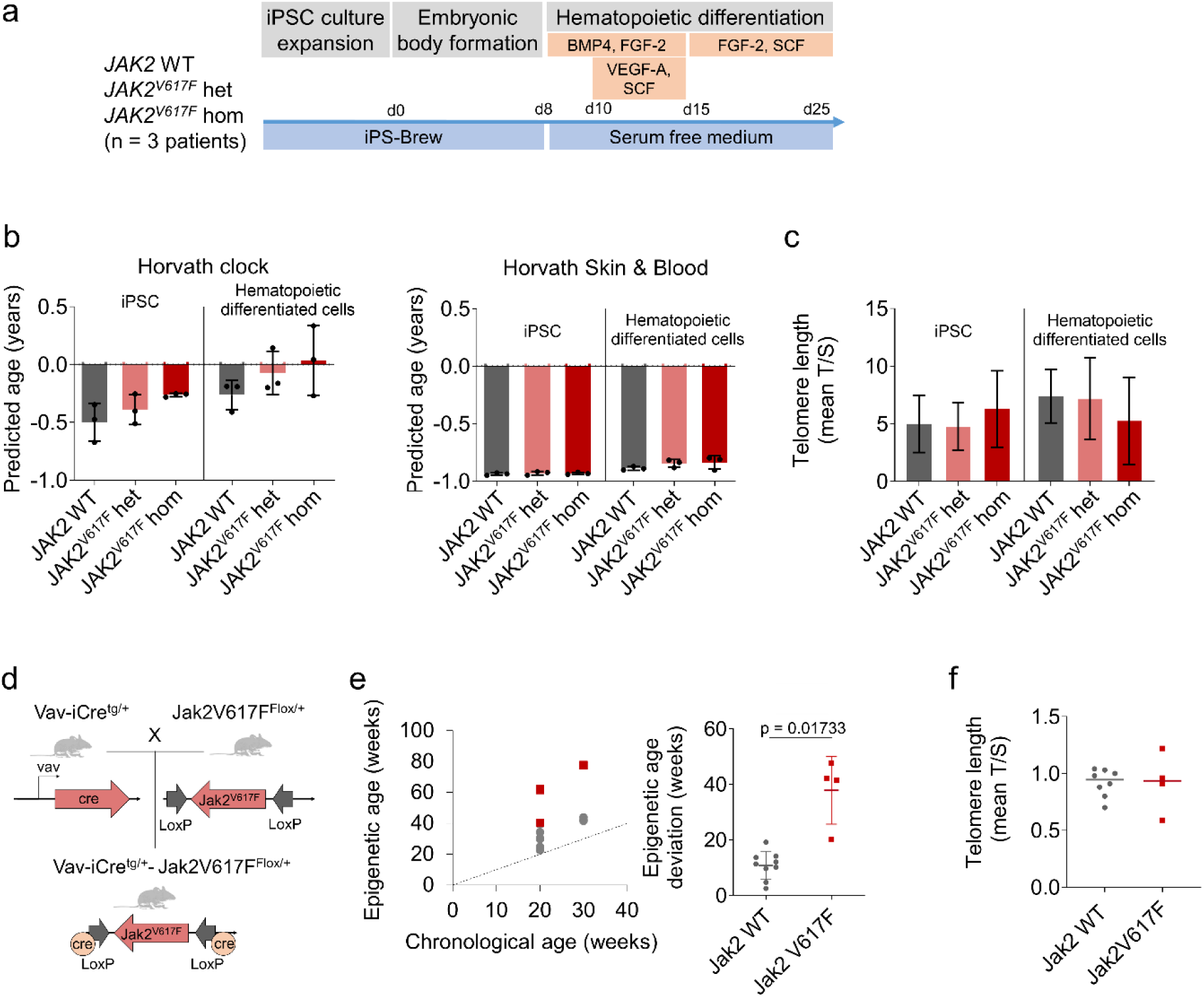
Model systems to investigate the effect of JAK2V617F mutation on cellular aging. **a)** Syngeneic iPSC lines with JAK2 WT, heterozygous, or homozygous JAK2V617F mutation were differentiated towards hematopoietic lineage. The differentiation protocol is indicated. **b)** Epigenetic age was estimated in iPSC and iPSC-derived hematopoietic cells base on Illumina BeadChip profiles using two alternative signatures: Horvath epigenetic clock (39) and Horvath Skin and Blood clock (40). **c)** TL measurement of iPSCs and iPSC-derived hematopoietic cells according to their genotype. **d)** Schematic representation of the Jak2V617F mouse model system. Vav-iCre transgenic (tg/+) mice were crossed with conditional knock out mice for Jak2V617F (flox/+) in the hematopoietic lineage. **e)** Epigenetic age in bone marrow derived cells from WT mice (grey) and Jak2V617F mice (red) is plotted against their chronological age (left). The epigenetic age deviation was compared between both groups (right, unpaired Welch’s t-test). **f)** Telomere length of WT versus Jak2V617F mice. colonies.

Next, we analyzed vav-cre driven *Jak2*^V617F^ transgenic mice after development of an MPN-like phenotype (Figure 5d) (27). When we analyzed epigenetic age in unfractionated bone marrow cells, there was a significant acceleration in mice with *Jak2*^V617F^ mutation (n = 4) as compared to WT *Jak2* (n = 9; Figure 5e, p = 0.02). TL did not differ between *Jak2*^WT^ and *Jak2*^V617F^ (Figure 5f), which might be due to different expression of telomerase and regulation of TL in mice (43).

### Effects of senolytic drugs on the malignant clones

Subsequently, we analyzed if cellular aging in MPN is also reflected by senescence-associated markers. In fact, senescence-associated genes were significantly enriched in MPN (Supplemental Figure S5a). Furthermore, we estimated the fraction of senescent cells by flow cytometric analysis of senescence-associated β-galactosidase activity. In PBMCs of MPN patients, this C12FDG staining was in tendency higher levels than in healthy controls (n = 7 and n = 6, respectively; p = 0.051; Supplemental Figure S5b). Overall, these results reinforced the conclusion that aspects of aging and cellular senescence are enhanced in MPN. Thus, particularly the malignant clones might be more susceptible to senolytic drugs than the remaining non-mutated cells.

To further address this question, we cultured PBMCs for three days with eight different drugs that have been suggested to specifically target senescent cells: piperlongumine, ABT-263, RG-7112, nutlin-3a, dasatinib in combination with quercetin, AMG-232, JQ1, and S63845. Furthermore, we tested the telomerase inhibitor BIBR-1532. After performing a concentration-dependent cell viability assay to determine the IC50 (Supplemental Figure S6), we treated PBMCs of 9 MPN patients with *JAK2*^V617F^ mutation with each compound using the concentrations below and above the IC50 (indicated as low and high concentration). After three days, the remaining *JAK2*^V617F^ mutation burden was analyzed with digital droplet PCR (ddPCR, Supplemental Figure S7). Overall, the senolytic drugs had only a moderate specific effect on the mutated subsets. Only JQ1, a potent inhibitor of the BET family of bromodomain proteins, and piperlongumine, an amide alkaloid constituent of the long pepper, showed a moderate but significant reduction of the *JAK2*^V617F^ allele burden (for piperlongumine only at low concentration; Figure 6a). In analogy, we analyzed if the treatment would also affect epigenetic age predictions (n = 7 for each compound). A moderate, non-significant reduction of epigenetic age-predictions was observed for RG7112 and again for piperlongumine (Figure 6b), potentially due to the depletion of clonal cells with premature epigenetic age. Furthermore, we measured TL with TEL-PCR (n = 5 for each compound). A significant increase in TL was again observed with JQ1, piperlongumine and nutlin-3a (Figure 6c). These results indicate that at least JQ1 and piperlongumine might have a selective effect for malignant cells with accelerated cellular aging.

**Figure 6.**
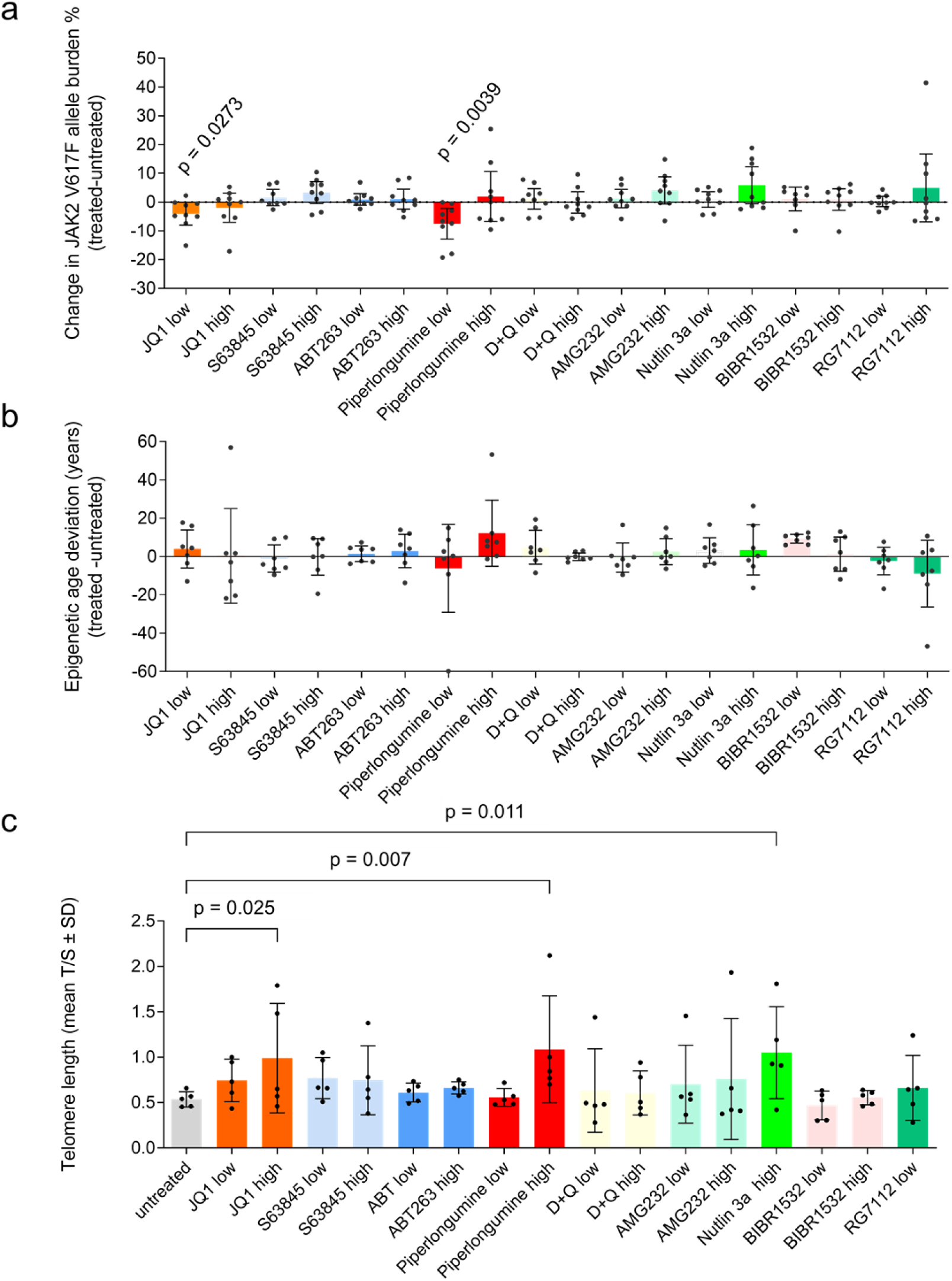
Testing of senolytic compounds on MPN samples in vitro. Peripheral blood mononuclear cells of MPN patients were cultured for three days with nine different compounds at either high or low concentration: JQ1 (10µM, 20µM), S63845 (500nM, 1µM), ABT-263 (100nM, 200nM), Piperlongumine (10µM, 50µM), Dasatinib in combination with Quercetin (D+Q; 20µM, 50µM), AMG-232 (1µM, 10µM), Nutlin-3a (10µM, 50µM), BIBR-1532 (50µM, 100µM), and RG-7112 (10µM, 50µM). **a)** *JAK2*^V617F^ mutational burden measured by ddPCR in untreated *versus* treated cells (n = 9; one-sample t-test). **b)** Epigenetic age changes in treated *versus* untreated cells (n = 7; measured by pyrosequencing; one-sample t-test). **c)** Changes in telomere length in treated *versus* untreated cells (n = 5; measured by TEL-PCR; one-way ANOVA). TL in Nutlin-3a 10 µM and RG7112 50 µM was not measurable due to a low concentration of DNA in the samples.

### The telomerase inhibitor BIBR-1532 induces senescence

BIBR-1532 is a potent telomerase inhibitor that has been tested previously in CML (28), but not yet systematically on BCR-ABL-negative MPN cells. Although short-term treatment with BIBR-1532 did not reduce the prominent malignant clone with accelerated aging, we hypothesize that inhibition of telomerase mediates proliferation-dependent critical telomere shortening, which eventually leads to telomere-mediated senescence or apoptosis. Therefore, an effect on subclones with particularly short TL might be observed only after longer treatment. To this end, we cultured PMF-derived PBMC (n = 9) in CFU assays with or without BIBR-1532, picked 30 colonies per condition, and analyzed the percentage of mutated colonies (*JAK2*^V617F^ and *CALR* rearrangements). Notably, the fraction of mutated colonies declined particularly in those patient samples that initially revealed shorter mean TL in granulocytes by flow-FISH analysis (R^2^ = 0.68; p = 0.0064; Figure 7a), indicating that samples with accelerated telomere attrition are more susceptible to telomerase inhibition. For five of these samples, we additionally performed NGS sequencing for the bulk of CFU colonies and the results substantiated that the effect of BIBR-1532 was more pronounced in samples with shorter telomeres, showing a reduction in mutant variant allele frequency only in patients with shorter telomeres (Supplemental Figure S8).

**Figure 7.**
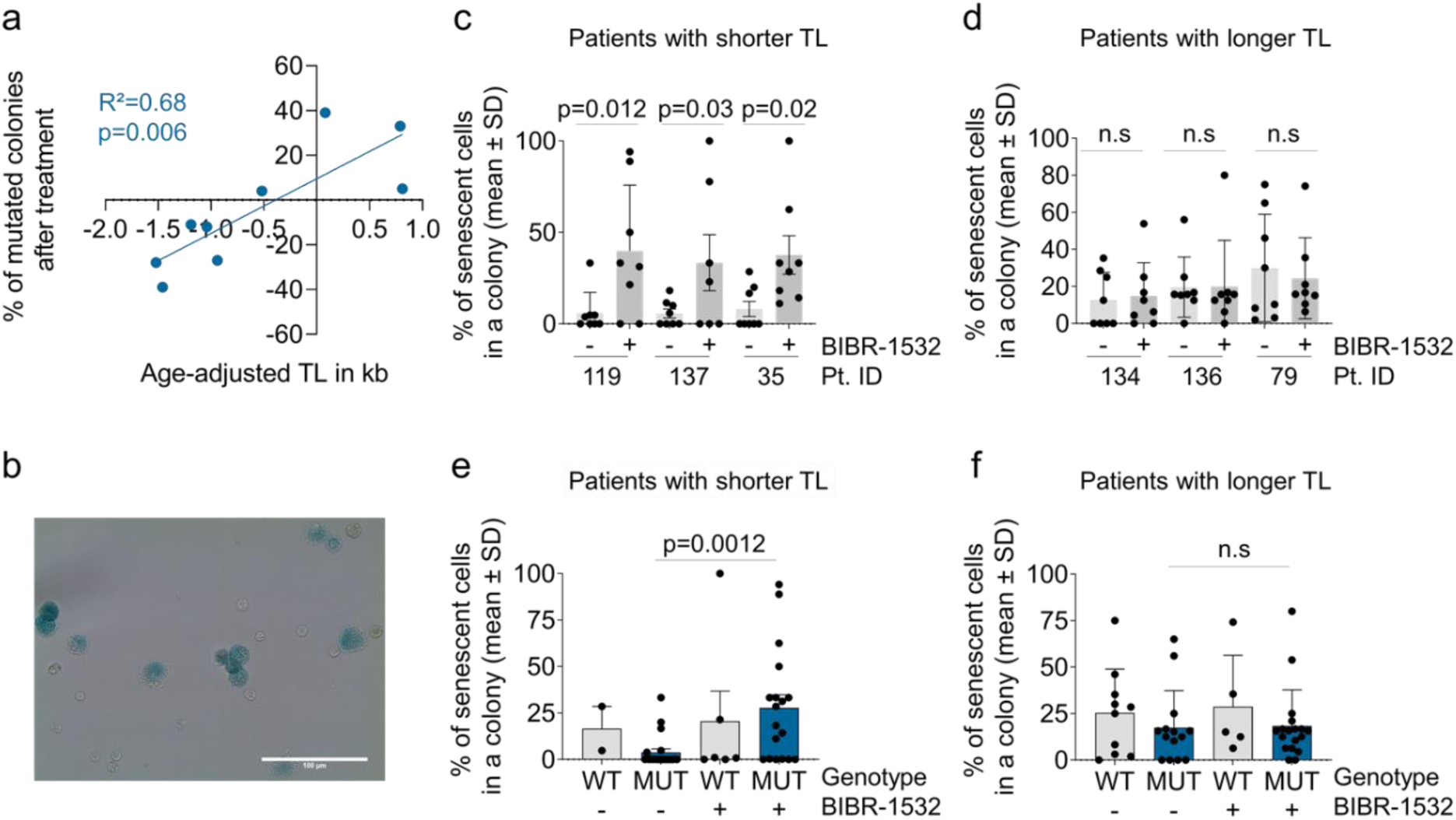
Telomerase inhibitor evokes senescence in samples with short telomeres. **a)** Peripheral blood mononuclear cells of nine PMF patients were seeded into a colony forming unit assay for 14 days either with BIBR-1532 (50 µM) or DMSO for control. The change in *JAK2*^V617F^ or *CALR* mutational burden was measured before and after treatment and correlated with the age-adjusted TL (measured by flow-FISH). **b)** ß-galactosidase staining within individual colonies depict senescent subsets. **c)** Percentage of senescent cells in eight colonies analyzed after treatment with BIBR-1532 (50 µM) or DMSO. Colonies were derived from patients with shorter telomeres (Patient# indicated). **d)** The same analysis was performed for patients with longer telomeres (paired t-student test). **e)** Percentage of senescent cells in WT and *JAK2*^V617F^ mutated or *CALR* rearranged colonies after treatment with BIBR-1532 (50 µM) or DMSO in the three patients analyzed in c. **f)** This analysis is also shown for the three patients of d (one-way ANOVA).

Subsequently, we performed ß-galactosidase staining within individual colonies (Figure 7b). In the three PMF donors with very short telomeres, the fraction of senescent cells clearly increased upon BIBR-1532 treatment (Figure 7c). In contrast, this effect was not observed in the three patients that revealed longer telomeres (Figure 7d). To further substantiate if the potential triggering of senescence occurs specifically in the malignant cells, we further analyzed the genotype in these colonies. BIBR-1532 treatment seems to specifically increase the fractions of senescent cells in CFUs carrying *JAK2*^V617F^ mutation or CALR rearrangements in patients with shorter telomeres (Figure 7e, f).

## Discussion

Cellular aging is heterogeneous in MPN samples and particularly accelerated in the mutant clone. Recently, McPherson and colleagues (29) used pyrosequencing of three age-associated CpGs in *ASPA, ITGA2B*, and *PDE4C* (26) to demonstrate a correlation between *JAK2* mutation allele frequency and epigenetic age. They observed accelerated epigenetic ageing only in PV, whereas we found this for all MPN entities. The highest epigenetic age acceleration was found in primary myelofibrosis, which might be attributed to expansion of the malignant clone, or additional mutations of this clinically advanced subtype (30). Furthermore, PMF revealed the most prominent telomere attrition in the granulocytic compartment, which is in line with a previous study showing pronounced TL attrition in both PV and PMF (31). We analyzed TL in both granulocytes and lymphocytes and a significant reduction of TL was only observed for granulocytes, in line with the restriction of MPN to the myeloid compartment (32). Notably, there was only a moderate correlation between TL attrition and epigenetic delta-age, which is in line with our previous observations on replicative senescence of mesenchymal stromal cells (33), blood of healthy donors (26), and disorder dyskeratosis congenita (34). Thus, telomere length and epigenetic age seem to reflect different biological properties of aging (35, 36).

Furthermore, our study demonstrates that both measures for cellular aging are accelerated in patient samples with higher *JAK2*^V617F^ allele frequency. Hence, it was conceivable that the *JAK2*^V617F^ mutation directly impacts on cellular aging. However, neither epigenetic age nor telomere attrition was clearly accelerated in iPSCs or iPSC-derived hematopoietic cells with *JAK2*^V617F^ mutation, as compared to the syngeneic wild type counterparts. It is conceivable, that the general reset of age-associated modifications during reprogramming into iPSCs (24, 26) masks some of the mutational effects, which would only become evident during longer differentiation. At least, *JAK2*^V617F^ does not directly accelerate cellular aging in this model system. On the other hand, the *Jak2*^V617F^ mouse model showed a prominent acceleration in epigenetic aging. While we previously hypothesized that chromatin reorganization in the course of epigenetic aging may contribute to acquisition of specific mutations (37), this does not seem to be the case here, because the *Jak2*^V617F^ mutation is driven by vav-cre and thus expressed in the entire hematopoietic system, resulting in enhanced epigenetic aging in this murine model system. Either way, it is remarkable that aspects of cellular aging were recapitulated in this murine model system, indicating that it may be a relevant aspect in the pathophysiology of disease progression.

Cellular aging is heterogeneous within a given sample (38) and we have previously demonstrated that this heterogeneity can be reflected by single-read age predictions in BA- seq datasets (19). MPN is a clonal disease and if the tumor-initiating cell reveals a distinct DNAm profile, it might be anticipated that this DNAm pattern remains prominent in the patient. However, single-strand epigenetic age-predictions were rather heterogeneous in the MPN samples and there was high variability at neighboring CpGs. This could be due to the very long development of MPN and the differences in clinical histories of the patient samples included here, which may entail increased epigenetic plasticity (39). In fact, when we analyzed age-related methylation in individual CFUs, which are notoriously single-cell derived within 14 days, this analysis revealed clearly prevalent DNAm patterns. Thus, our analysis can provide some insight into the dynamics of DNAm changes, which are captured by the CFU initiating cell during 14 days of culture expansion but diversified over decades of MPN development.

When focusing on telomere length in PV, we observed significantly shorter telomeres in *JAK2*- mutated colonies when compared to their WT counterpart, suggesting that accelerated aging is specifically restricted to the malignant clones. This is further supported by the finding that the WT colonies of PV patients revealed similar TL as colonies of healthy donors. Notably, in colonies of PMF patients, we did not observe a difference in terms of TL between mutant and WT colonies derived from the same patient. This might be attributed to accumulating cell extrinsic factors in the hematopoietic stem cell niche of the more progressive PMF that may play a role by disfavoring normal hematopoiesis (40).

Treatment with senolytic drugs is a new concept that raises high hopes to support healthy aging (41, 42). While cellular aging and senescence are not identical, the two processes are clearly interlinked. In fact, we found evidence that also senescence-associated features, such as gene expression signatures and β-galactosidase are enhanced in MPN. In our exploratory analysis of potentially senolytic compounds we observed a moderate reduction of *JAK2*^V617F^ allele burden and increased average TL upon treatment with JQ1 and piperlongumine. These substances might therefore have some specificity for the malignant subset particularly for subclones with shorter telomeres. JQ1 is a BET inhibitor that induces apoptosis in tumor cells by downregulating E2f/p21 signaling and regulating c-MYC, and it has also been used in clinical trials in patients with AML and myelodysplastic syndromes (43). Notably, Kleppe and colleagues demonstrated a reduction in disease burden of MPN by JQ1 treatment either alone or in combination with ruxolitinib (44). Piperlongumine is a potent cytotoxic component, which selectively induces ROS production via inhibition of antioxidant enzymes (45). Furthermore, piperlongumine was shown to target senescent human WI-38 fibroblasts (46) and eliminate cancer cells by blocking the JAK2-STAT3 pathway (47). Taken together, these senolytic drugs may specifically target the malignant clone in MPN. However, it must be considered that the compounds may also have non-senolytic effects, particularly at these relatively high concentrations during short-term *in vitro* treatment.

Finally, our data indicate that the telomerase inhibitor BIBR-1532 might be particularly effective on PMF samples with more pronounced telomere attrition. For these patients, we observed a clear reduction of CFUs carrying either *JAK2*^V617F^ or *CALR* rearrangements. This exemplifies that TL measurement might become an important parameter and potentially predictive biomarker for personalized medicine in MPN. Furthermore, our results indicate that BIBR-1532 might induce senescence, particularly in the mutant compartment of MPN. Cells with shorter TL are already more susceptible to chromosomal damage (48) and targeting telomerase with TI was shown to disrupt telomere stability and restore susceptibility through senescence (49). While BIBR-1532 has so far not been evaluated in clinical trials, telomerase inhibition with imetelstat has already demonstrated clinical activity in primary, post-ET- or post-PV myelofibrosis (15–17). Another study on non-small cell lung cancer cell lines demonstrated that particularly those lines with short TL were susceptible to imetelstat treatment (50). It will therefore be important to determine if particularly patients with short telomeres at diagnosis profit from imetelstat treatment and if this telomerase inhibitor might also induce senescence in the mutant compartment of MPN. It might even be conceivable to combine TI and senolytics to accelerate senescence specifically in the malignant stem cell clones in MPN patients (by the TI component) and to finally eradicate it (with a senolytic drug).

## Data Availability

The datasets used and/or analysed in this current study are available from the corresponding author on reasonable request.

## Acknowledgements

We thank Anne Abels for technical assistance and Kim Kricheldorf for organizing the patient samples, Isabelle Plo and the Institut Gustave-Roussy, Villejuif, France, for kindly providing the *Jak2*^V617F^ transgenic mice, and Caroline Küstermann and Janik Böhnke for support in iPSC culture. This work was supported by the Flow Cytometry Facility, a core facility of the Interdisciplinary Center for Clinical Research (IZKF) Aachen within the Faculty of Medicine at RWTH Aachen University. We thank Lichterzellen (www.lichterzellen.de) and AA/PNH e.V. for their initiative, enthusiasm and continued support to T.H.B and M.V.; M.A.S.T was funded by CAPES-Alexander von Humboldt postdoctoral fellowship (99999.001703/2014-05) and donation by U. Lehmann. This work was supported by funds from the German Research Foundation (Deutsche Forschungsgemeinschaft, DFG) within CRU344 “Untangling and Targeting Mechanisms of Myelofibrosis in Myeloproliferative Neoplasms” (WA 1706/12-1 and WA1706/14-1; KO2155/7-1; BR1782/5-1; ZE432/10-1) and KO2155/6-1; by Deutsche Krebshilfe (TRACK-AML); and the ForTra gGmbH für Forschungstransfer der Else Kröner-Fresenius-Stiftung.

## Authors’ contributions

M.V., V.T., and M.Ka. performed experiments and analyzed the results. M.N. performed BA-seq analysis. M.S. supported cell culture and sequencing experiments. M.Ki. analyzed NGS analysis of mutational burden. J.B. supported gene expression analysis and the murine model system. N.F., M.A.S.T., and M.Z. generated and characterized the *JAK2*^V617F^ iPSC lines. S.K. interpreted data and supported analysis. T.H.B., F.B., and W.W. conceptually designed the study and supervised the research. M.V., V.T., and W.W. wrote the first draft of manuscript. All authors read and approved the final manuscript.

