## Supplemental Figures and Tables for "Cellular aging is accelerated in the malignant clone of myeloproliferative neoplasms"

### Supplemental Methods

#### Analysis of mutational burden

A clinically validated amplicon-based next-generation sequencing (NGS) panel (Truseq Custom Amplicon Kit, Illumina, San Diego, USA) was used to analyze the coding region of 32 genes that typically harbor mutations associated with hematologic malignancies, as described previously (1, 2). Variants were reviewed manually, using a bidirectional frequency of >1% for driver mutations and > 5% for additional mutations as cutoffs. Alternatively, the JAK2$^{V617F}$ allele burden was analyzed with digital droplet PCR (ddPCR) using the mutation assay dHsaMDV27944642 from Bio-Rad, according to the manufacturer's instructions.

#### Telomere length measurements

Flow-fluorescent *in situ* hybridization (flow-FISH) (3, 4): Cryopreserved whole blood cells after red blood cell depletion were used for the flow-FISH analysis of telomere length (TL) in lymphocytes and granulocytes, as previously described (4). Briefly, samples were prepared for cell denaturation and mixed with a telomere-specific (CCCTAA)3-peptide nucleic acid FISH probe labeled with FITC (Eurogentec, Liège, Belgium) for DNA hybridization. DNA counterstaining was performed with LDS 751 (Sigma-Aldrich, Missouri, USA). Granulocytes and lymphocytes are stained by LDS 751 and can be distinguished by signal intensities in both the FL-3 channel as well as in the forward scatter (FSC). TL of bovine thymocytes was determined by Western blot (19.515 kb) and used as a reference to convert the TL of granulocytes and lymphocytes into kb. An FC 500 flow cytometer (Becton Dickinson, East Rutherford, USA) was used for data acquisition. All measurements were carried out in triplicates. Healthy controls (n = 134) were used for age adaptation of TL.

Telomere PCR (TEL-PCR) (5): This method was used for TL measurement in colony forming units (CFUs), CD34$^{+}$ cells, iPSCs before and after hematopoietic differentiation and in mouse bone marrow-derived cells. 1.4 ng of genomic DNA per reaction was used in the Absolute Human TL Quantification qPCR Assay Kit (ScienCell, Carlsbad, USA) and FastStart Essential DNA Green Master (Roche, Basel, Switzerland). TL measurements are given in T/S ratios. A T/S ratio is calculated by dividing the number of copies of the telomere template (T) by the single copy reference (SCR) template (S), which is an amplified 100 bp region on human chromosome 17. The TL q-RT-PCR was performed according to the manufacturer's instructions.

#### Epigenetic age prediction with targeted bisulfite amplicon sequencing

Genomic DNA was isolated from cryopreserved whole blood cells after red blood cells depletion with the QIAamp DNA Mini Kit (Qiagen, Hilden, Germany), or for colonies in the CFU assay with NucleoSpin XS Tissue Kit (Macherey-Nagel, Düren, Germany). DNA was quantified with a Nanodrop 2000 Spectrophotometer (Thermo Scientific, Wilmington, USA) and bisulfite converted with the EZ DNA Methylation Kit (Zymo Research, Irvine, USA). For targeted bisulfite amplicon sequencing (BA-seq) three age-associated CG dinucleotides (CpG sites) that are associated with the coiled-coil domain-containing protein 102B (*CCDC102B*), four and a half LIM domains protein 2 (*FHL2*), and phosphodiesterase 4C (*PDE4C*) (6) were amplified by PyroMark PCR kit (Qiagen) using primers with handle sequences for the subsequent barcoding step, as described in detail before (6). PCR conditions are summarized in Supplemental Table S1. The three amplicons of each donor were pooled at equal concentrations, quantified with Qubit (Invitrogen, Massachusetts, USA), and cleaned up with paramagnetic beads from Agencourt AMPure PCR Purification system (Beckman Coulter, California, USA). Four microliters of PCR products were subsequently added to 21 µl PyroMark Master Mix (Qiagen) containing 0.4 µM of barcoded primers (adapted from NEXTflexTM 16S V1-V3 Amplicon Seq Kit, Bioo Scientific, Austin, USA) for a second PCR. PCR products were again quantified with the Qubit, combined in equimolar ratios, and cleaned by Select-a-Size DNA Clean & Concentrator Kit (Zymo Research). A 12-pM DNA library was diluted with 15% PhiX spike-in control and eventually subjected to 250 bp pair-end sequencing on a MiSeq lane using the Miseq reagent V2 Nanokit (both from Illumina).

FastQ files from MiSeq analysis were aligned to the reference genome *hg19* using the Bismark tool (7) and DNA methylation values determined with the Bismark methylation extractor. For heatmaps, the frequencies of DNA methylation patterns in individual reads were calculated by the number of reads containing the pattern divided by the total number of reads of the target region per sample. The most abundant reads of similar patterns within neighboring CpGs were grouped for visualization with Python's package seaborn (8). Epigenetic age was calculated as follows:

Predicted age (in years) = 3.86 + 0.825 $DNAm^{FHL2}$ - 0.342 $DNAm^{CCDC102B}$ + 1.177 $DNAm^{PDE4C}$

Alternatively, epigenetic age predictions were based on the binary sequel of methylated and non-methylated CpGs within individual reads of BBA-seq data, as described in our previous study (6).

**Epigenetic age prediction with pyrosequencing**

Pyrosequencing was used for fast and robust epigenetic age predictions of mouse bone marrow cell pellets and peripheral blood mononuclear cells (PBMCs) from patients after senolytic drug treatment *in vitro*. For mouse epigenetic age, three age-associated CpG sites were measured in Proline rich membrane anchor 1 (*Prima1*), Heat shock transcription factor 4 (*Hsf4*) and Potassium voltage-gated channel modifier subfamily S member 1 (*Kcns1*), as described in our previous work (9) and updated by selecting neighboring CpGs for the Prima1 region. Epigenetic age was calculated as follows:

Predicted age (in weeks) = - 6.325 - 0.308 $DNAm^{Prima1}$ + 2.588 $DNAm^{Hsf4}$ + 1.003 $DNAm^{Kcns1}$

For pyrosequencing analysis of the samples that were treated with senolytic compounds we analyzed for DNAm changes at the same age-associated CpGs of *PDE4C*, *CCDC102B* and *FHL2*, as described in detail before (6).

**Magnetic separation of CD34+ cells**

PBMCs of MPN patients were isolated by gradient centrifugation and enriched for $CD34^+$ cells by magnetic-activated cell sorting (MACS) with microbeads (Miltenyi Biotech, Germany). DNA was isolated before and after $CD34^+$ enrichment with the Monarch® PCR & DNA Cleanup Kit (New England Biolabs, Frankfurt, Germany) and then used for TL measurements.

**Senescence associated beta-galactosidase (SA-ß-gal) assay**

As a surrogate marker for senescence, we stained CFUs for β-galactosidase (Cell Signaling Technology, Danvers, USA). Single colonies were seeded on individual poly-L-lysine coated glass slide (Merck KGaA, Darmstadt, Germany) and stained according to the manufacturer's instructions. Cells were observed under a microscope (Leica DMRX, Leica Microsystems, Wetzlar, Germany).

Alternatively, we used fluorescent based assay for quantification of senescence phenotype in both MPN patients (n=7) and healthy donors (n=6; one outlier was removed). For that, PBMCs were pre-treated with 100nM bafilomycin A1 (medchemexpress) for one hour and then treated with 33 µM 5-dodecanoylaminofluorescein di-β-D-galactopyranoside (C12FDG, Abcam) for 2 hours at 37°C (10). Cells were collected, washed with PBS and stained with anti-CD34-APC (BD) at a dilution of 1:100 for 30 minutes at 4°C. Cells were washed with PBS + 2% FCS and measured by flow cytometry using a FACS Canto II (BD). Relative ß-gal activity was estimated based on the setting of gates for FSC *versus* SSC, excluding dead cells and cellular debris and also the population of CD34+ senescent cells were analyzed using FlowJo.

**Analysis of genes associated with senescence in the microarray data**

Gene expression profiles from 6 healthy donors, 6 ET, 11 PV and 9 PMF patients were published by Baumeister, J *et al.* in GSE174060 (11) and used for senescence pathway analysis (12). Significantly differentially regulated genes were selected by a Benjamini–Hochberg adjusted p value <0.05 and log2-fold changes above 0.5 or below −0.5. Enrichment of senescence-associated genes was estimated with hyper geometric distribution analysis.

**Generation and hematopoietic differentiation of $JAK2^{V617F}$ iPSC**

Induced pluripotent stem cells (iPSC) from three PV patients were generated by reprogramming PBMCs as described before (13, 14) (Flosdorf *et al.*, manuscript submitted). In patient 1 with 37% $JAK2^{V617F}$ allele burden in the PBMCs, only WT (Human Pluripotent Stem Cell Registry; UKAi002-A) and heterozygous (UKAi002-B) iPSC clones were obtained after reprogramming. Therefore, CRISPR/Cas9 genome engineering was used to introduce the $JAK2^{V617F}$ mutation generating a homozygous $JAK2^{V617F}$ iPSC clone (UKAi002-B3). Similarly, in patient 2 with 96% $JAK2^{V617F}$ allele burden in the PBMCs gave rise to only homozygous iPSC clones (UKAi003-A), and the heterozygous (UKAi003-A2) and wild type ones (UKAi003-A1) were generated by CRISPR/Cas9 repair. In patient 3 with 25% $JAK2^{V617F}$ allele burden in the PBMCs, only WT (UKAi016-A) and heterozygous (UKAi016-B) clones were obtained, and CRISPR/Cas9 was used to generate homozygous clones (UKAi016-B1).

Hematopoietic differentiation of iPSC clones was performed as described previously (15). In brief, iPSCs were cultured in micro-contact printed plates with StemMACS iPS Brew XF medium (Miltenyi Biotec) and 10 µM Y-27632 (Abcam, Cambridge, United Kingdom) to form embryonic bodies (EB) (16). Self-detachment of EBs was observed after 6 to 9 days, depending on the clone, and they were then resuspended in serum-free medium containing 50% IMDM, 50% Ham's F12, 1% chemically defined lipid concentrate, 2 mM GlutaMAX (all Thermo Fisher Scientific), 0.5% Albiomin (Unifols), 400 µM 1-

thioglycerol, 50 µg/mL L-ascorbic acid, and 6 µg/mL holo transferrin (all Sigma Aldrich, St. Louis, MO, USA) supplemented with 10 ng/mL FGF-2 (Peprotech, Hamburg, Germany) and 10 ng/mL BMP-4 (Miltenyi Biotec). Approximately 30 to 50 EBs were distributed per well on a gelatin coated 6-well plate. From day 2 to day 7, cells were cultured in serum-free medium supplemented with 10 ng/mL FGF 2, 10 ng/mL BMP-4, 50 ng/mL SCF, 10 ng/mL VEGF-A (all Peprotech), and 10 U/mL penicillin/streptomycin (Thermo Fisher Scientific). From day 8 to day 16, serum-free medium was supplemented with 10 ng/mL FGF 2 and 50 ng/mL SCF only. Cells were harvested on day 16 and their phenotype was analyzed by flow cytometry and colony forming assay. For details on the hematopoietic differentiation and characterization we refer to our previous work (15).

**DNA methylation analysis using BeadChip data**

For iPSC and iPSC-derived hematopoietic cells the above-mentioned targeted signatures for epigenetic age-prediction could not be applied, because they were specifically trained for primary hematopoietic cells. Thus, we used multi-tissue epigenetic age predictors for Illumina BeadChip data. Genomic DNA was isolated from the 9 iPSC clones and iPSC-derived hematopoietic differentiated cells with the QIAamp DNA Mini Kit (Qiagen, Hilden, Germany), and bisulfite converted and hybridized with the Illumina human EPIC methylation microarray at Life and Brain (Life and Brain GmbH, Bonn, Germany). Data was processed using the Sesame package for R (17), normalized and corrected for dye bias, background, and mask probes with poor design. Additionally, CpGs at the X and Y chromosomes were removed and also the probes with failed detection P values, with resulting in 630,700 CpGs. Epigenetic age was predicted using Horvath's clock (18) and Skin and blood Horvath clock (19) using watermelon package for R (20).

**Jak2$^{V617F}$ mouse model**

The Vav-Cre-lox system was used to induce the *Jak2*$^{V617F}$ mutation in the mouse model, as described previously (21). Six wild type (WT) and three heterozygous *Jak2*$^{V617F}$ (VF) mice at 20 weeks of age and 3 WT and 1 VF mice at 30 weeks of age were used for epigenetic age prediction and telomere length measurements. The genotype of the mouse was confirmed by PCR, and massive splenomegaly and elevated myeloid and erythroid markers were observed in VF mice compared with WT mice. Bone marrow cells were collected for analysis by flushing tibiae and femurs, and red blood cells were lysed.

**Testing of senolytic drugs and a telomerase inhibitor**

Our choice of eight different senolytics was based on the different apoptotic mechanisms and the involvement of anti-apoptotic proteins, which have been shown to induce apoptosis in senescence cells. JQ1 is a BET inhibitor that targets non-homologous end joining to eliminate senescent cells, which is relevant in repairing the double stand breaks in DNA and activating the autophagy pathway (22). BH3-only BCL-2 family proteins are effectors of canonical mitochondrial apoptosis that have pro-apoptotic functions through BH1–3 pro-apoptotic proteins, such as BAX and BAK, while their activity is suppressed by BH1–4 anti-apoptotic BCL-2 family members (23). Here we used the first clinically approved BH3-mimetic ABT263 (Navitoclax) that target multiple BCL-2 proteins such as BCL-2, BCLxL and BCL-w (24), and S63845 that selectively targets the anti-apoptotic protein MCL-1 (25). Dasatinib is a tyrosine kinase inhibitor, which interferes with EFNB- dependent suppression of apoptosis. Quercetin has been shown to interfere with several anti-apoptotic pathways (26). Combination treatment showed a reduction in p16 and p21 expressing cells and SA-ß-gal positive cells (27). MDM2 inhibitors, which target the interaction of MDM2 with p53 and reactivate functional p53, lead the cell death of senescent cells (28). Depending on the binding site of p53-MDM2, different structurally unique small inhibitors were selected, such as AMG232, which is used in clinical trials in MPN patients who failed JAK inhibitor treatment (NCT03662126), RG7112, which is also used in clinical trials of leukemia patients (NCT01970930), and Nutlin-3a, which increases the degree of apoptosis in MPN by increasing p53 and p21 protein levels (29). Piperlongumine is a natural product that selectively kills cancer cells by inhibiting oxidative stress response proteins which are important for cancer cell survival in the presence of elevated ROS levels (30).

To estimate the senolytic activity of these compounds on cellular subsets with $JAK2^{V617F}$ mutation, we cultured cells for three days at different concentrations and analyzed proliferation and viability. PBMCs of MPN patients were cultured in StemSpan Serum-Free Expansion Medium (Stemcell Technologies, Vancouver, Canada), supplemented with 10 ng/mL SCF, 20 ng/mL TPO, 10 ng/mL FGF-1 (all PeproTech, Hamburg, Germany), 10 µg/mL heparin (Ratiopharm, Ulm, Germany), and 100 U/mL penicillin/streptomycin (Lonza, Basel, Switzerland) for three days. Selected nine drugs (eight senolytic compounds and the telomerase inhibitor BIBR-1532) were tested for their impacts on cell viability after three days with Cell Titer-Glo 2.0 luminescent cell viability assay (Promega, Wisconsin, USA) in 96-well plates that were seeded with 10 000 cells/well (3 wells per condition), using BioTek Synergy 2 plate reader and Gen5 software (Agilent Technologies, California, USA).

For a comparative approach of these compounds, we estimated the half-maximal inhibitory concentration (IC50) in our culture setting and used one concentration above and one below IC50 (diluted water or DMSO, according to the manufacturer’s instructions): nutlin3a (10µM (31) and 50µM, Selleck Chemicals LLC, Munich, Germany), JQ1 (10µM (32), 20µM, Sigma-Aldrich), ABT263 (100nM, 200nM (33), Selleck Chem), piperlongumine (10µM (34), 50µM, Selleck Chem), S63845 (500nM, 1µM (35), Selleck Chem), RG7112 (10µM (36), 50µM, Selleck Chem), dasatinib combined with quercetin (20µM (37), 50µM, both), AMG232 (1µM (38), 10µM, Axon medchem, Groningen, Netherlands), and the telomerase inhibitor BIBR-1532 (50µM (39), 100µM, Selleck Chem). Alternatively, cells were cultured in a 24-well plate seeded with 250 000 cells/well (2 wells per condition) for DNA isolation. The effect of BIBR-1532 on the clonogenic potential was tested at a concentration of 50 µM in the CFU medium during 14 days of culture.

While the drug concentrations used in this study may appear relatively high, particularly with regard to long-term treatment or *in vivo* applications, they were at a similar range as described by other studies (Nutlin 3a, 10 µM (31); JQ1, 1-10 µM (32); ABT-263, <1µM (33); Piperlongumine, 10 µM (34); S63845, 5 nM -1 µM (35); RG 7112, 2.5- 5 µM (36); Dasatinib (D) + Quercetin (Q), 20 µM D + 15 µM Q, (40); AMG-232, 1 µM (38); and BIBR 1532, 50 µM (39)). However, at these concentrations some of the compounds may have additional effects beyond senolytics.

#### Statistics

Linear regressions, mean absolute deviation (MAD), and mean absolute error (MAE) of age-predictions were calculated with Excel. Statistical analysis was performed with GraphPad Prism using one sample t-test, unpaired t-test paired t-test or one-way ANOVA. P values ≤0.05 were considered as indicative of statistical significance. IC50 values were calculated by nonlinear regression analysis using GraphPad Prism.

### Supplemental Figures

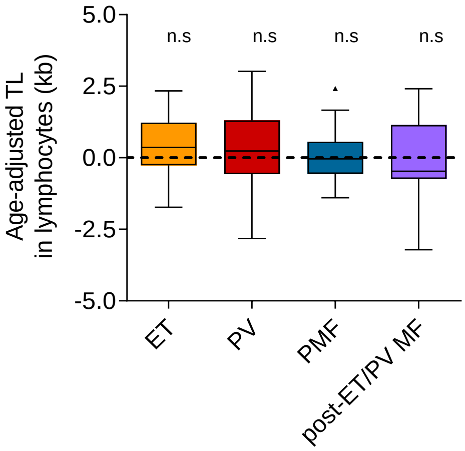

**Figure S1: Age-adjusted telomere length in lymphocytes in different MPN entities**
Telomere length (TL, in kb) was measured in lymphocytes via flow-FISH in blood of MPN patients (n = 129). One-sample t-test was used to calculate statistical significance. There is no significant reduction in telomere length in the lymphocyte compartment of any MPN entity.

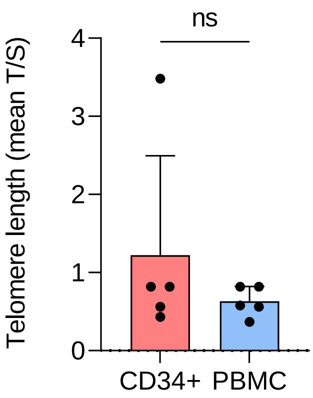

**Figure S2: Telomere length in CD34⁺ cells of MPN patients**
Telomere length (TL) was measured in MACS-sorted PB-derived $CD34^+$ cells and in PBMC via TEL-PCR of 5 MPN patients (4 PV, 1 post-PV-MF). A paired t-test was used to calculate statistical significance. These results are in line with previous reports that demonstrated correlation of TL in $CD34^+$ cells with TL in granulocytes or other leukocyte subsets in healthy donors (41).

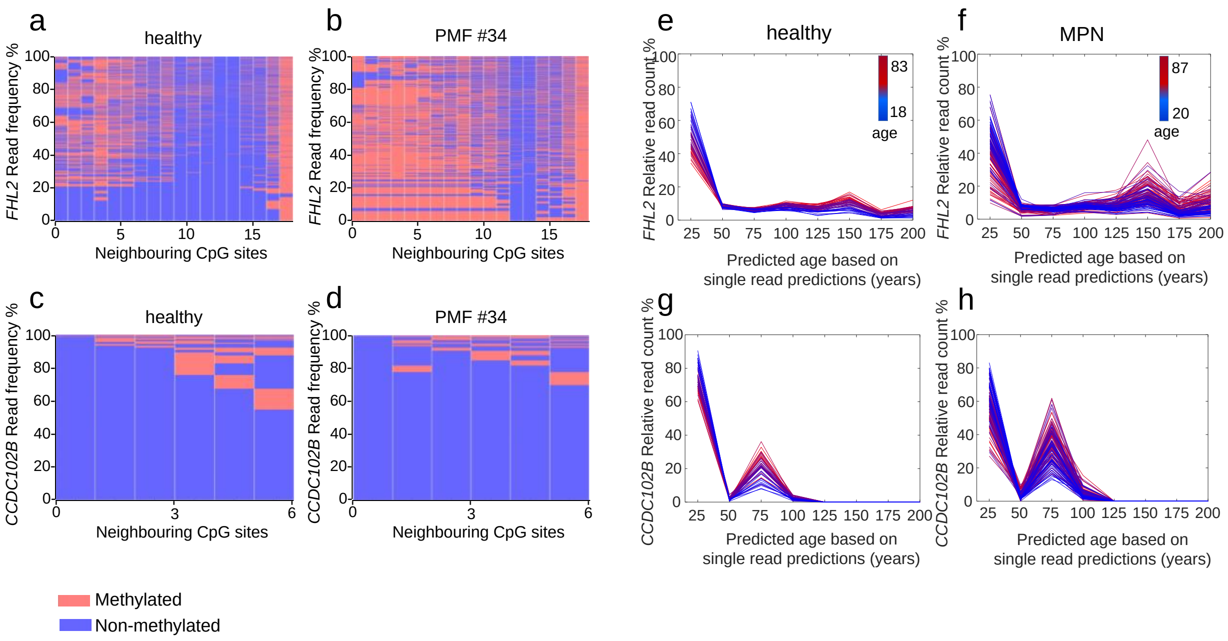

**Figure S3: Heterogeneity of epigenetic age predictions based on individual amplicon reads**
**a,b)** Heat map depicts exemplarily the frequencies of DNAm patterns within the neighboring CpGs of the *FHL2* amplicon in BBA-seq data of a) a healthy donor, and b) a PMF patient of the same age (in analogy to figure 3a,b). **c,d)** Furthermore, exemplary heatmaps are also shown for the *CCDC102B* amplicon. In healthy and MPN samples the patterns of neighboring CpGs are quite heterogeneous and there is no obvious clonal DNAm pattern in the patient samples. **e-h)** The binary code of methylated and non-methylated CpGs of each amplicon was then used for single read prediction for the e.f) *FHL2* amplicon, and the g,h) *CCDC102B* amplicon (in analogy to figure 3 c,d). The histograms demonstrate the heterogeneity of single read predictions (ranging from 0 to 200 years), and their relative frequency are provided for all healthy donors (18 to 83 years old) and MPN patients (20 to 87 years old). As expected, the higher predictions increase in elderly donors. Notably, the heterogeneity of epigenetic age predictions increases particularly in elderly MPN patients.

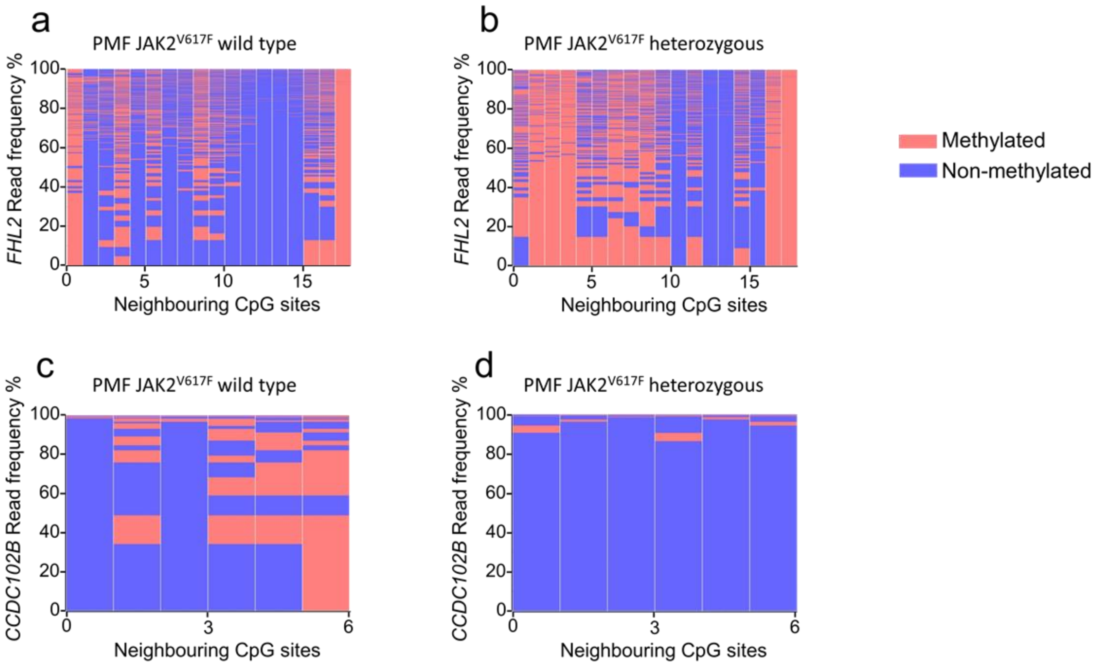

**Figure S4: Age-associated DNA methylation is more homogenous in colony forming units** Individual colony forming units (CFUs) were analyzed after 14 days (in analogy to figure 4b). a,b) The heat maps exemplarily depict frequencies of DNAm patterns within the neighboring CpGs of the *FHL2* amplicon in BBA-seq data of **a)** WT, and **b)** JAK2$^{V617F}$ mutated colonies from the same patient. **c,d)** In analogy, we analyzed the amplicons of *CCDC102B* for c) WT, and d) JAK2$^{V617F}$ colonies from the same patient. In comparison to the corresponding heatmaps of blood (Figure S1 a-d) there are consistently more prominent patterns in individual colonies, which might reflect the pattern of the initial colony forming unit.

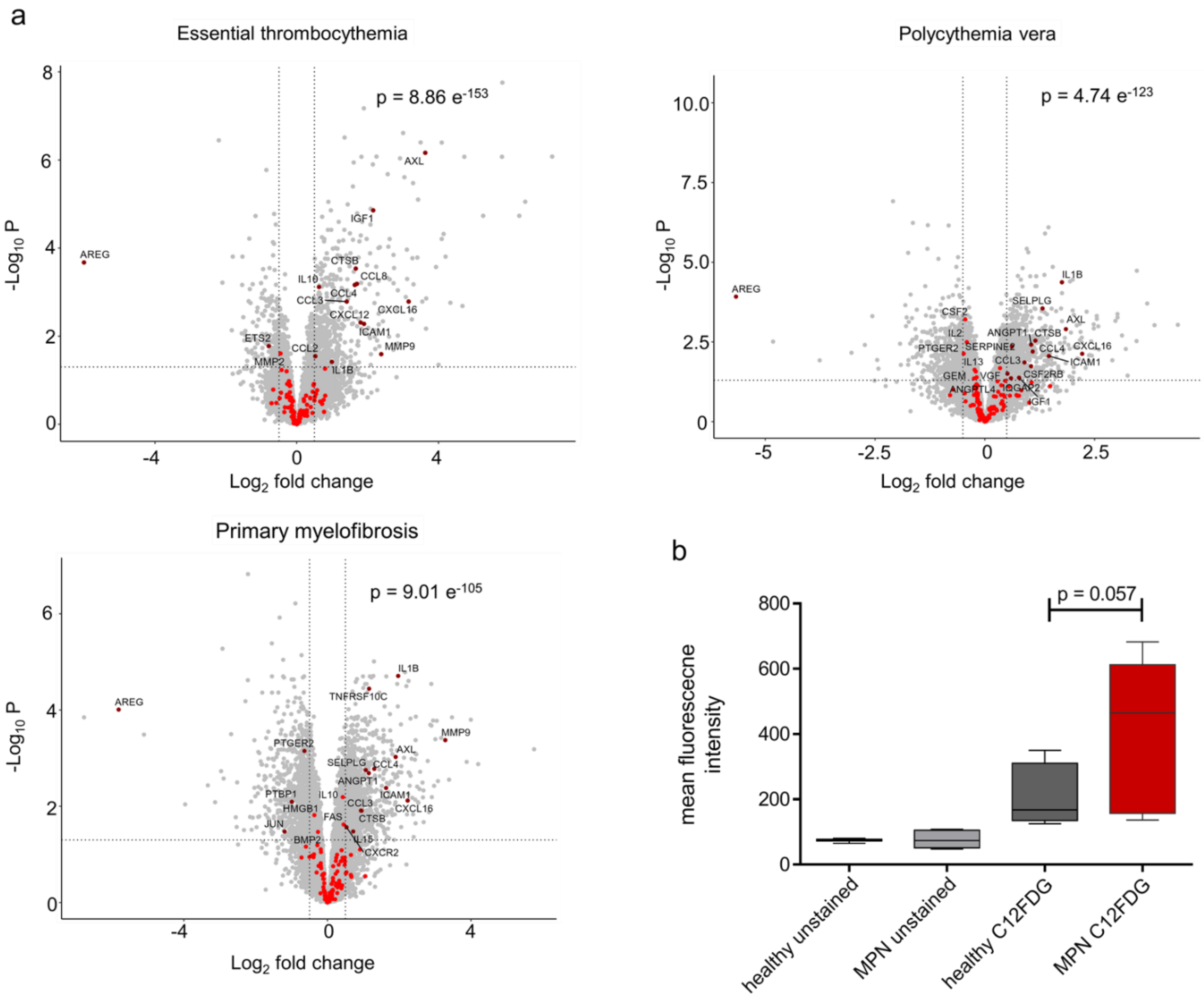

**Figure S5: Senescence phenotype is increased in MPN**

**a)** Gene expression profiles of 6 ET, 11 PV and 9 PMF patients, each compared with 6 healthy donors (25) (GEO submission number GSE174060) and were analysed for senescence-associated gene expression signatures. To this end, we focused on a set of 125 genes (SenMayo) that are differentially expressed during senescence (12). Significantly differentially regulated genes were selected by a Benjamini–Hochberg adjusted p value <0.05 and log2-fold changes above 0.5 or below −0.5. In fact, differential gene expression in MPN samples *versus* healthy donors revealed significant enrichment of this senescence-associated gene set for all MPN entities (p-value estimated by hyper geometric distribution). **b)** PBMC of MPN patients (n = 7) and healthy donors (n = 6; one outlier was removed) were stained with C12FDG and analyzed by flow cytometry. The mean fluorescence intensity was in tendency higher in MPN samples, indicating that they possess higher β-galactosidase activity.

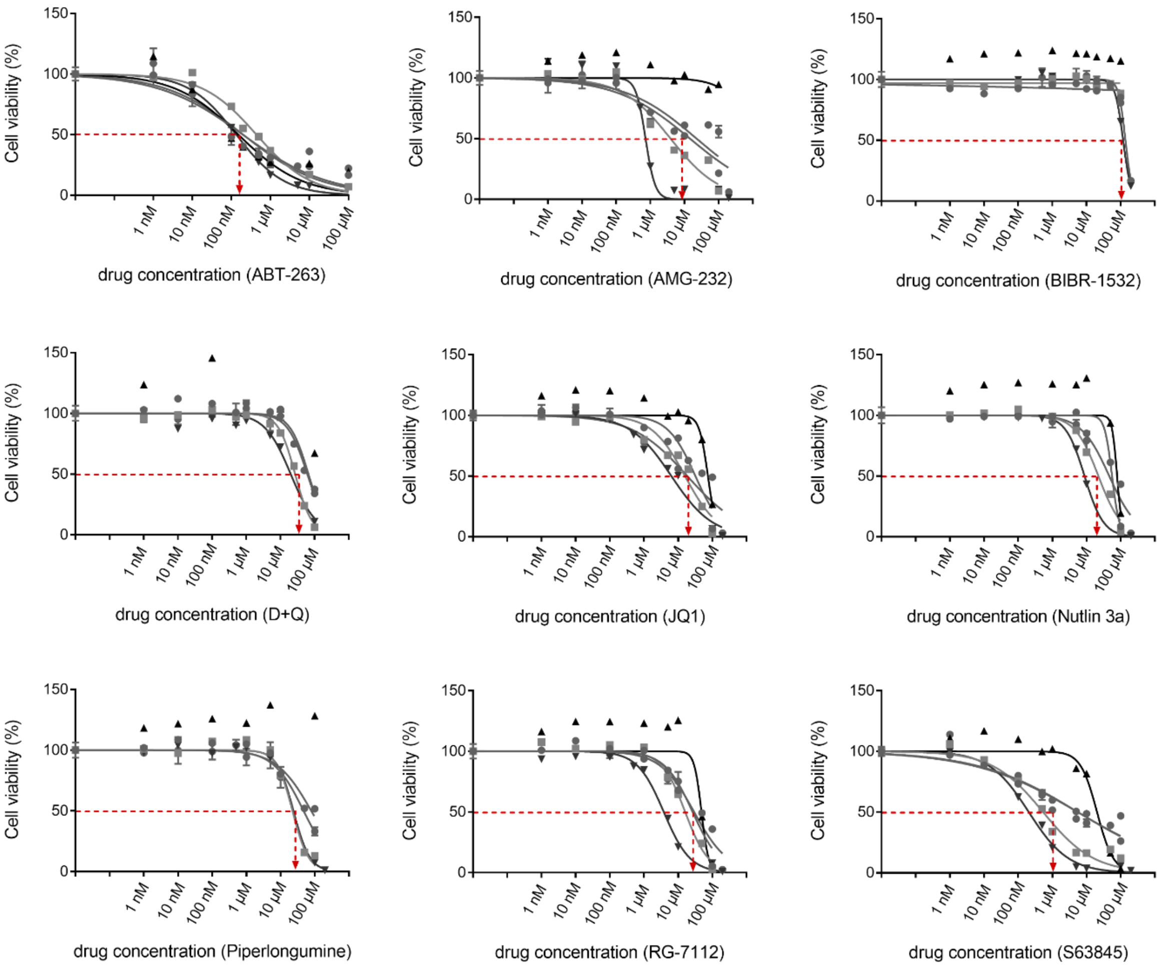

**Figure S6: Concentration dependent viability upon senolytic drug treatment**
To determine the dose range for subsequent analysis of senolytic compounds, PBMCs from MPN patients (79, 132, 135, 146 and 147) were cultured for three days in 96-well plates at different concentrations as indicated. Each curve represents a patient sample, which was performed in triplicate. Viability was then determined using CellTiter Glo assay and median IC50 values were calculated for each drug; ABT-263: 196.9 nM, AMG-232: 9.2 µM, BIBR 1532 128.3 µM, D+Q: 44.15 µM, JQ1: 18.6 µM, nutlin 3a: 46.4 µM, piperlongumine: 35.5 µM, RG7112: 26.7 µM, S63845: 5.5 µM.

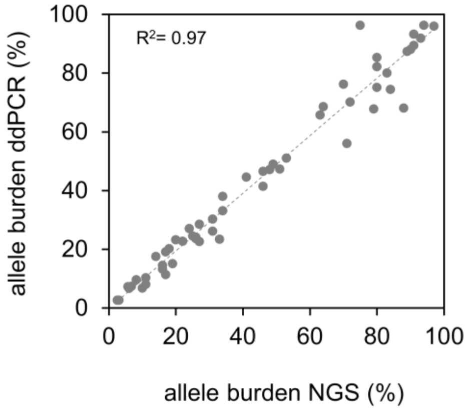

**Figure S7: Analysis of JAK2$^{V617F}$ mutation burden with digital droplet PCR *versus* deep sequencing**
The fraction of JAK2$^{V617F}$ mutations was analyzed in PBMCs of 50 MPN patients. Both methods show a very good correlation.

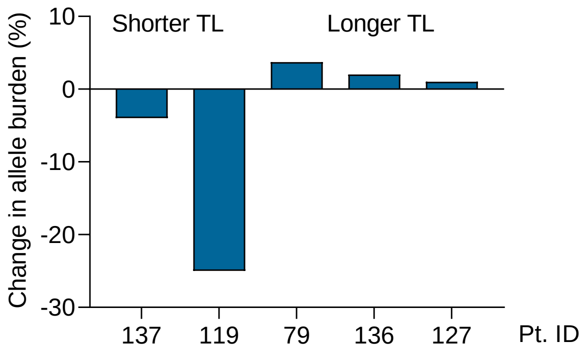

**Figure S8: Next generation sequencing data of JAK2$^{V617F}$ mutations upon BIBR 1532 treatment**
The JAK2$^{V617F}$ mutational burden (%) was analyzed with NGS in colonies after telomerase inhibition with BIBR 1532 as compared to non-treated controls. In PMF patients that revealed shorter telomeres at the time of diagnosis the CFUs revealed a stronger effect of BIBR 1532 treatment.

### Supplemental Tables

**Table S1: PCR conditions for bisulfite barcoded amplicon sequencing**

| Step | PCR1 (*CCDC102B* and *FHL2*) | | | PCR1 (*PDE4C*) | | | PCR2 | | |
|---|---|---|---|---|---|---|---|---|---|
| | **Temp.** | **Time** | **Cycles** | **Temp.** | **Time** | **Cycles** | **Temp.** | **Time** | **Cycles** |
| Enzyme activation | 95 °C | 15 min | | 95 °C | 15 min | | 95 °C | 15 min | |
| Denaturation | 95 °C | 30 sec | 40x | 95 °C | 3 sec | 35x | 95 °C | 30 sec | 16x |
| Annealing | 56 °C | 30 sec | | 53 °C | 35 sec | | 60 °C | 30 sec | |
| Extension | 72 °C | 30 sec | | 72 °C | 35 sec | | 72 °C | 30 sec | |
| Final extension | 72 °C | 10 min | | 72 °C | 10 min | | 72 °C | 10 min | |
| Hold | 4 °C | ∞ | | 4 °C | ∞ | | 4 °C | ∞ | |

1.5mM of $MgCl_2$ was used in both PCRs. These same conditions are used for all AML-associated regions.

**Table S2: Primer list for the genotyping of single CFU colonies**

| Primer | Sequence |
|---|---|
| CALRdel52 Frw | ACAACTTCCTCATCACCAACG |
| CALRdel31 Rev | GGCCTCAGTCCAGCCCTG |
| CALRins5 common Frw | TAACTGCAGTGTCAGCGGTG |
| CALRins5 non-mutated allele Rev | TGTCCTCATCATCCTCCTTG |
| CALRins5 mutant Rev | TGTCCTCATCATCCTCCGAC |
| JAK2V617F Frw | TCCTCAGAACGTTGATGGCAG |
| JAK2V617F Rev | GTTTTACTTACTCTCGTCTCCACAAAA |
| JAK2 WT Frw | GCATTTGGTTTTAAATTATGGAGTATATG |
| JAK2 Rev | ATTGCTTTCCTTTTTCACAAGAT |

Abbreviations: Frw: forward; Rev: reverse
